# Joint human and animal vaccination strategies to reach mobile pastoralists and their livestock in Africa: A scoping review

**DOI:** 10.1101/2025.09.26.25336600

**Authors:** Majdi M. Sabahelzain, Adeline Tinessia, Catherine King, Rebika Nepali, Siddig Mohamedali, Janeth George, Victoria J. Brookes, Kerrie Wiley

## Abstract

**Background:** Africa’s pastoralist population is approximately 268 million, which is over a quarter of the continent’s population. They occupy more than 40% of Africa’s land, though density varies by country. Mobile pastoralists are often underreported in national censuses due to political marginalisation and difficulties in counting these groups. Vaccination services delivered through fixed facilities do not adequately reach mobile pastoralists, which impacts vaccination coverage and health outcomes for these mobile groups who are economically and culturally connected to their livestock. This scoping review explored literature on joint human-animal vaccination strategies as a One Health approach to improve vaccination coverage among these mobile pastoralists and their animals.

**Methods:** A systematic search was performed across 12 databases, following the Joanna Briggs Institute (JBI) methodology and adhering to the PRISMA-ScR reporting guidelines.

**Results:** Of 2469 records screened for eligibility, 17 articles were included in a full-text assessment, and five met the eligibility criteria. The included studies were published between 2004 and 2016. Three studies were conducted in Chad and one each in Nigeria and Somalia. The implementation of the vaccination campaigns was organised in three phases: planning, implementation, and post-vaccination. Key enabling factors were identified in these joint campaigns, including the use of a participatory approach throughout both the planning and implementation phases, with a strong emphasis on intersectoral collaboration and active community engagement in all activities. Interventions were adapted during the campaigns to ensure they were contextually acceptable. The joint human and animal vaccination campaigns increased vaccination coverage among mobile pastoralists compared to baseline levels by targeting children, women of reproductive age, and their animals. Nevertheless, the campaigns highlighted human vaccination results but paid little attention to animal populations, contradicting the values of mobile pastoralists.

**Conclusions:** This review offers valuable insights into the design of joint vaccination campaigns tailored for mobile pastoralists and their livestock. Whilst joint vaccination campaigns appear to improve human vaccine uptake, more information is needed to evaluate the potential benefits that could be gained by attention to animal vaccination and improved animal health outcomes, which could further improve non-vaccine-related human health outcomes.

## 1. Introduction

The pastoralist population in Africa is estimated to be 268 million, representing over a quarter of the continent’s total population [1]. Little (2015) defines pastoralism as ‘‘the practice of keeping livestock as a means of either primary or secondary subsistence’’ [2]. The degree of pastoralism and mobility varies among different groups, i.e. pastoralism exists on a continuum. It encompasses practices that range from a combination of plant cultivation and animal herding (agropastoralism) to a nearly exclusive focus on herding. Movement patterns can differ significantly, from sedentary utilisation of fertile pasturelands (with minimal migration) to transhumance (seasonal migration), seminomadic (permanent homesteading), and fully nomadic pastoralism (characterised by frequent movements of homesteads) [2].

Although pastoralists live in areas that cover more than 40% of Africa’s landmass, and it is known that the population density and distribution vary significantly between countries [1], mobile pastoralists in Africa, especially the nomadic population, are not well captured in national censuses and surveys [3]. This stems from various factors including political and administrative marginalisation, and reluctance to engage with the government, driven by motives like tax evasion or cultural beliefs, such as concerns that strangers might attract the ‘evil eye’—belief that certain people, often strangers or envious individuals, can harm others, their children, livestock, or possessions just by looking at them with jealousy or ill intent will [3–5]. This can make it challenging to create effective policies to address the unique needs of these communities [3].

Vaccine-preventable diseases rank as the primary cause of death among children under five in Sub-Saharan Africa [6]. Research into polio outbreaks has shown that a significant number of wild poliovirus cases in Chad, Nigeria, and Somalia, including the final cases in Chad and Somalia, occurred among nomads [7–9]. Vaccination coverage among nomadic and settled children shows a significant disparity. In Chad, only 7% of children in pastoralist communities receive the Bacillus Calmette–Guérin (BCG) vaccine, compared to 79% in settled areas. Polio vaccination is similarly low at 11% for pastoralists, while settled communities achieve 80% coverage [10]. In Kenya, around 60% of nomadic children are unvaccinated, versus 7.2% in pastoralist settlements [11]. In addition, nomadic populations in Africa face a heightened risk of various diseases [12], including zoonoses such as brucellosis and Rift Valley fever (RVF), due to their close interactions with livestock [13]. These zoonoses cause significant health challenges and economic burdens in endemic regions due to regular contact with livestock and frequent cross-border travel. Despite the public health risks, targeted policies or interventions to improve vaccination access for nomadic pastoralists in Africa are lacking [14–16].

The formal vaccination delivery system traditionally relies on fixed sites, which are not always accessible to mobile pastoralist communities [14,16]. Fixed vaccination sites are generally designed for settled populations, making them less accessible to mobile pastoralist communities whose lifestyles and remote locations prevent regular access to centralised facilities [14]. Vaccination strategies using mobile teams, which could potentially reach these populations more effectively, have been underutilised [14–16]. This has led to limited improvements in vaccination coverage and health outcomes among mobile pastoralists. However, this delivery system was designed to prioritise humans, and in many pastoralist communities, including nomadic communities in Africa in which identity is linked to cultural values around livestock and mobility, animals are prioritised due to the deep connection of people to their animals [17]. Therefore, the western-style structure of human health service delivery is not valued as highly in these communities [18,19]. This is demonstrated by previous research, which has found that livestock vaccination rates in these nomadic communities often surpass those for humans [20]. It is known that successful health interventions often rely on cultural sensitivity, community engagement, and service mobility [14,21]; therefore, we hypothesise that joint vaccination approaches for humans and animals could improve uptake for both.

Joint vaccination approaches for humans and animals in pastoralist communities, which live at the ecosystem-animal-human interface, embody the One Health concept, recognising the interconnectedness of human and animal health and the environment [22]. This integrated approach is particularly relevant for nomadic pastoralist communities with typically limited access to primary health care for either humans or animals, yet for whom zoonotic diseases pose significant threats at the human-animal interface and climate and ecosystem-related diseases pose threats for both animals and their people. By simultaneously addressing both human and animal health needs, joint vaccination campaigns could create synergistic benefits such as reducing the transmission of zoonotic pathogens like anthrax between species, which are critically important to pastoralist communities [23], leveraging limited healthcare resources in remote areas, and respecting the non-hierarchical worldview of pastoralists who value animal health alongside human health [24]. For the pastoralist communities, livestock represents their primary source of sustenance and is essential for their economic well-being and social status, making the health of their animals critically important [17]. In many of these communities, traditional healers also provide care for both humans and animals, highlighting that animal health is a vital entry point for the provision of both human and animal health understanding and services [17,25,26].

Although the One Health approach has been criticised for being overly reductionist [27] and having yet to address the absence of hierarchical considerations for certain populations (including Indigenous and nomadic pastoralists), the added value of applying this approach can outweigh its limitations [18,28]. For example, in resource-limited settings such as nomadic pastoral communities, it has proved to increase financial savings through intersectoral cost sharing [29]. In Kenya, One Health has also been shown to improve vaccine coverage by enhancing cost-effective resource utilisation [30] and fostering community trust through multi-level stakeholder engagement in Chad [20].

Vaccine service delivery models tailored for non-hierarchical health world-views, such as those held by some nomadic populations, are needed to simultaneously promote human and animal health. The objective of this scoping review was to identify and explore non-hierarchical models for delivering health services to pastoralist communities, with a focus on co-delivering human and animal vaccinations to reach mobile pastoralists and their livestock in Africa. This aligns with the growing emphasis on the One Health approach across Sub-Saharan Africa [31] and provides a comprehensive assessment and synthesis of these interventions.

## 2. Methods

This scoping review was conducted according to the Joanna Briggs Institute (JBI) guidelines [32]. Additionally, we consulted the Preferred Reporting Items for Systematic Reviews and Meta-Analyses extension for Scoping Reviews (PRISMA-ScR) checklist [33] for reporting.

### Search strategy

A systematic search of key bibliographic databases, including OVID Medline (1946-August 08, 2025), OVID Embase Classic (1947-1973), OVID Embase (1974-August 082025), OVID Global Health (1910-Week 32, 2025), SCOPUS (1788-18 August, 2025), African Index Medicus (1993 – August 2025) and Web of Science Core Collection including Science Citation Index Expanded (1900-August 2025), Social Sciences Citation Index (1900-August 2025), Arts & Humanities Citation Index (1975-August 2025), Emerging Sources Citation Index (2005-August 2025), Conference Proceedings Citation Index-Science (1990-August 2025) and Conference Proceedings Citation Index-Social Science & Humanities (1990-August 2025) was undertaken by an experienced information specialist (CK). We searched key websites, including those of WHO, UNICEF, the Global Polio Eradication Initiative (GPEI), FAO, the Technical Network for Strengthening Immunization Services (TechNet-21), and Google Scholar, for relevant grey literature.

The search terms were intentionally broad due to the lack of standardised language or medical subject heading (MeSH) terms related to pastoralists and animals or livestock. Search terms were informed by content experts’ and information specialists’ input, as well as analysis of terms used in previous reviews.

The search strategy, including all identified keywords and index terms, was adapted to incorporate the syntax of each included database and/or information source. The Ovid Medline search strategy is available in (Supplementary 1). Reference lists of all included sources of evidence were screened for additional studies.

### Study selection

Studies were included in this review if they met the following criteria: the study designs were primary research, the population encompassed patterns of movement of pastoralism (specifically transhumance, seminomadic or nomadic), the setting was confined to the African continent, the strategies involved vaccinating both the pastoralists and their animals, and there were no restrictions on outcomes, date of publication, or language. Studies were excluded from this review if they were not primary research, the population consisted only of non-pastoralists or sedentary pastoralists, the setting was outside Africa, or the strategies focused on vaccinating only pastoralists or only their animals.

After the initial search, all identified citations were uploaded into EndNote, and duplicates were removed. Two or more independent reviewers screened the titles and abstracts using Covidence to assess them against the review’s inclusion criteria (MS, AT, RN, SM). Potentially relevant sources were retrieved in full. The full text of selected citations was thoroughly evaluated by two or more independent reviewers against the inclusion criteria (MS, AT, RN, SM). Any full-text sources of evidence that did not meet the inclusion criteria were recorded and reported as exclusions. Any disagreements between the reviewers at each stage of the selection process were resolved through discussion or with a third reviewer (KW). Full-text articles not in English were translated using Google Translate’s document feature to allow for screening and data extraction.

### Data extraction

Data were extracted from papers included in the scoping review by two or more independent reviewers (MS, AT, RN) using a data extraction tool developed and piloted by the reviewers. The following data from the studies were extracted; study title, name of lead author, year of publication, country in which the study conducted, study aim, study design, setting, methods used, study duration, population description (human and animal), description of the strategy used, comparison, description of the strategy and intervention, information on human and animal vaccines in the study, disease outbreak, the outcome expected, measured and observed in human and animals, and limitations as noted by the author and limitations as noted by the reviewers.

### Data analysis and presentation

The extracted data were charted and summarised narratively.

## 3. Results

### 3.1. Screening

Figure 1 shows a PRISMA flow chart of articles included and excluded in each stage of identification and screening. The database searches identified 2,469 records. After removing duplicates, 1,141 records were screened for eligibility based on their title and abstracts. After 1,124 were excluded, 17 were included for full-text screening. Records were most often excluded because they were not primary research or were not interventional research. Overall, five records were included for data charting and synthesis. Any screening discrepancies were resolved via discussion.

**Figure 1:**
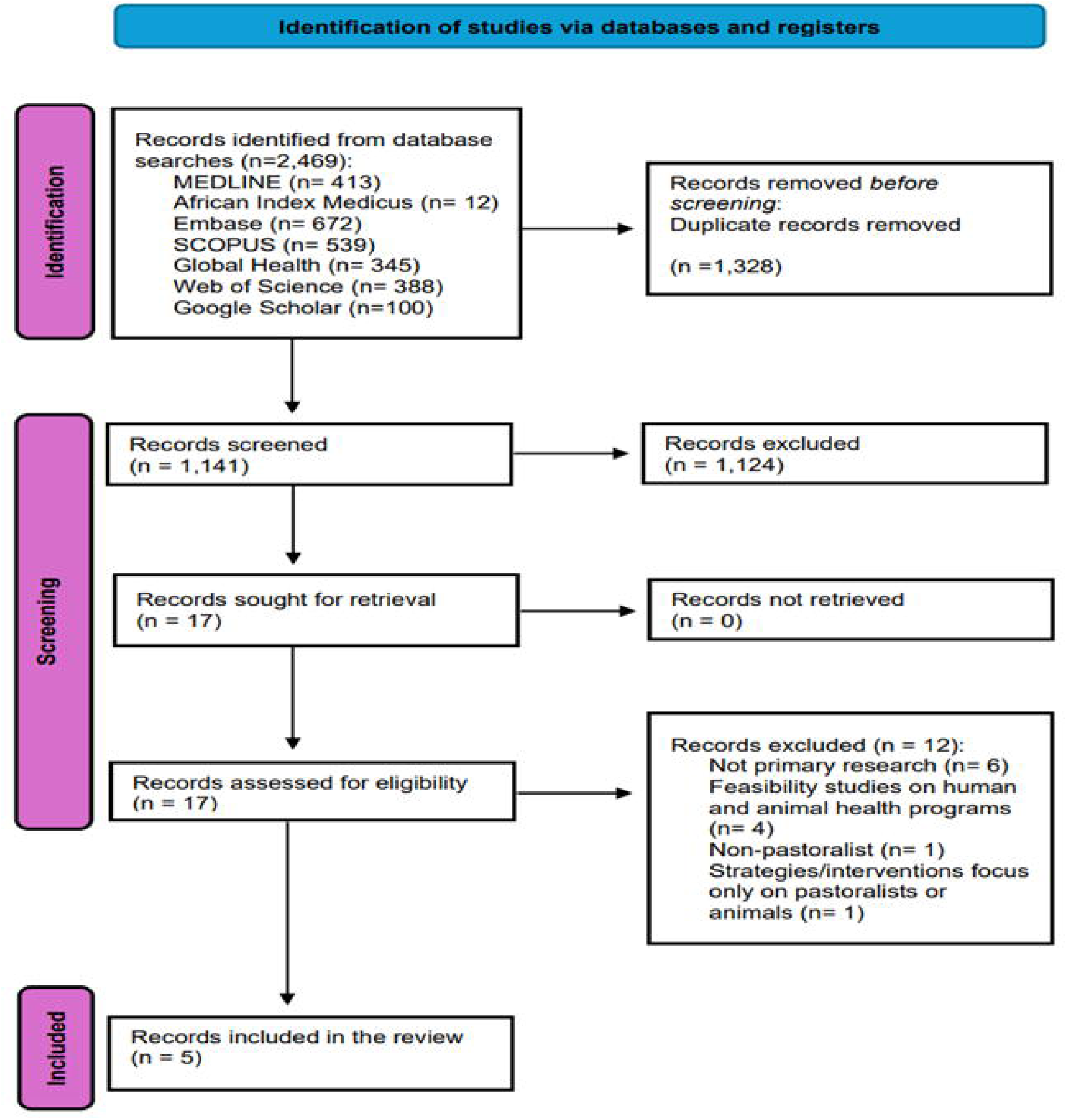
PRISMA flowchart of the results of the record screening and selection process.

### 3.2. Data Charting

One record was published in a peer-reviewed conference proceedings [34], while the remaining records were published in four peer-reviewed journals. All records were published between 2004 and 2016. Three studies were conducted in Chad [35,36,37], one in Nigeria [34], and one in Somalia [38]. Four records were published in English, and one was published in French [36].

Table 1 summarises the location by country, funding sources, the study type and aim, methodologies, a description of human and animal populations, and expected and measured outcomes for human and animal vaccinations.

**Table 1.**
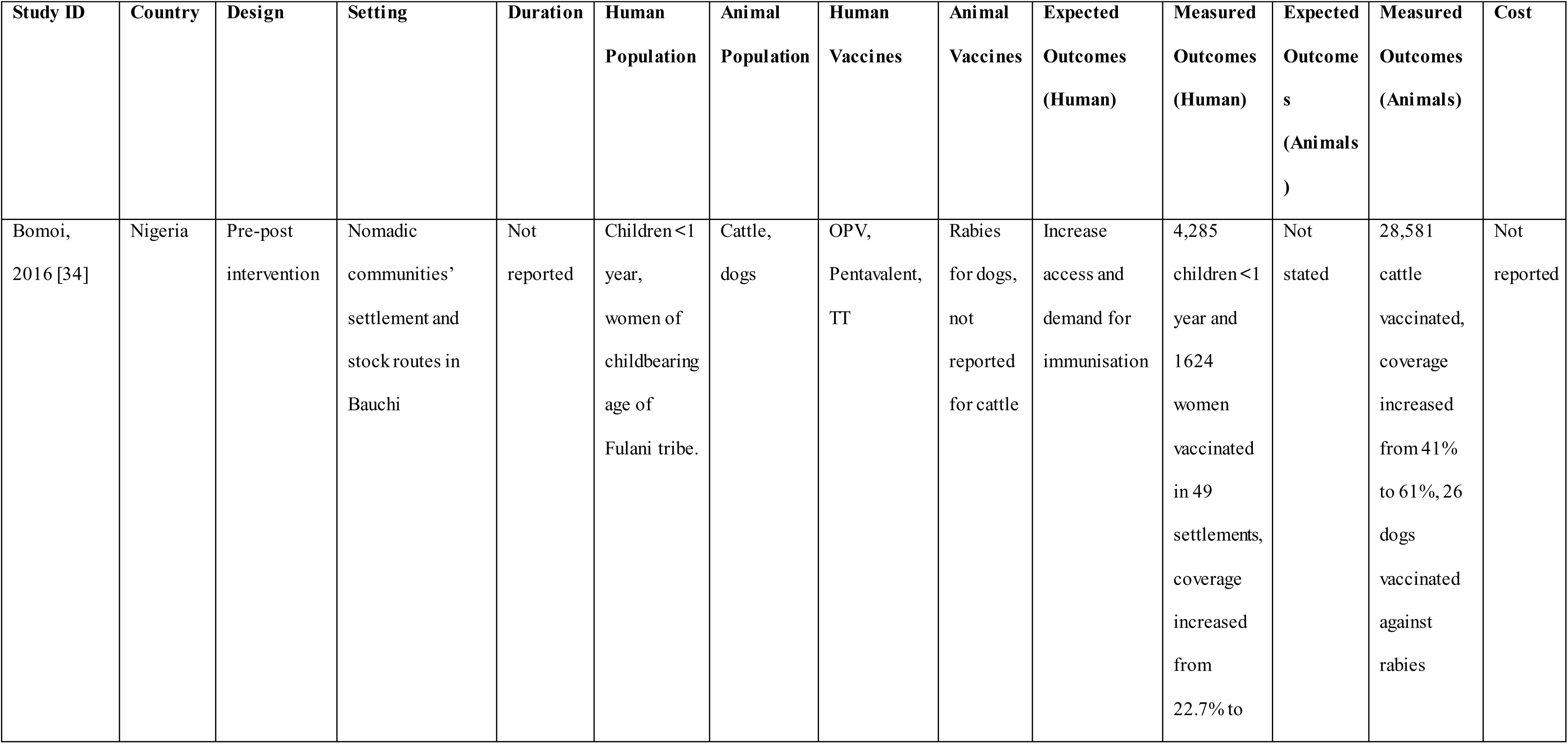
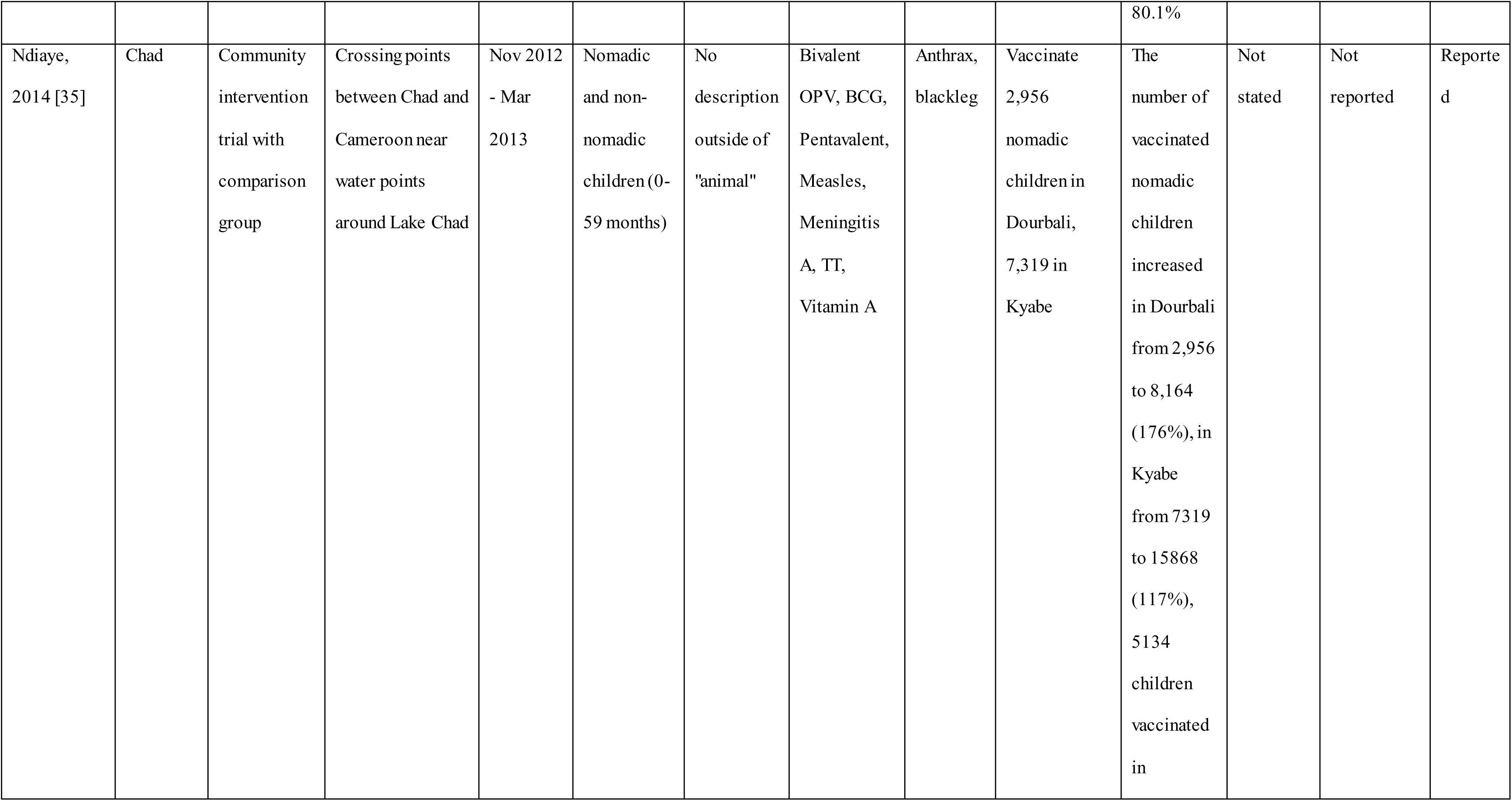
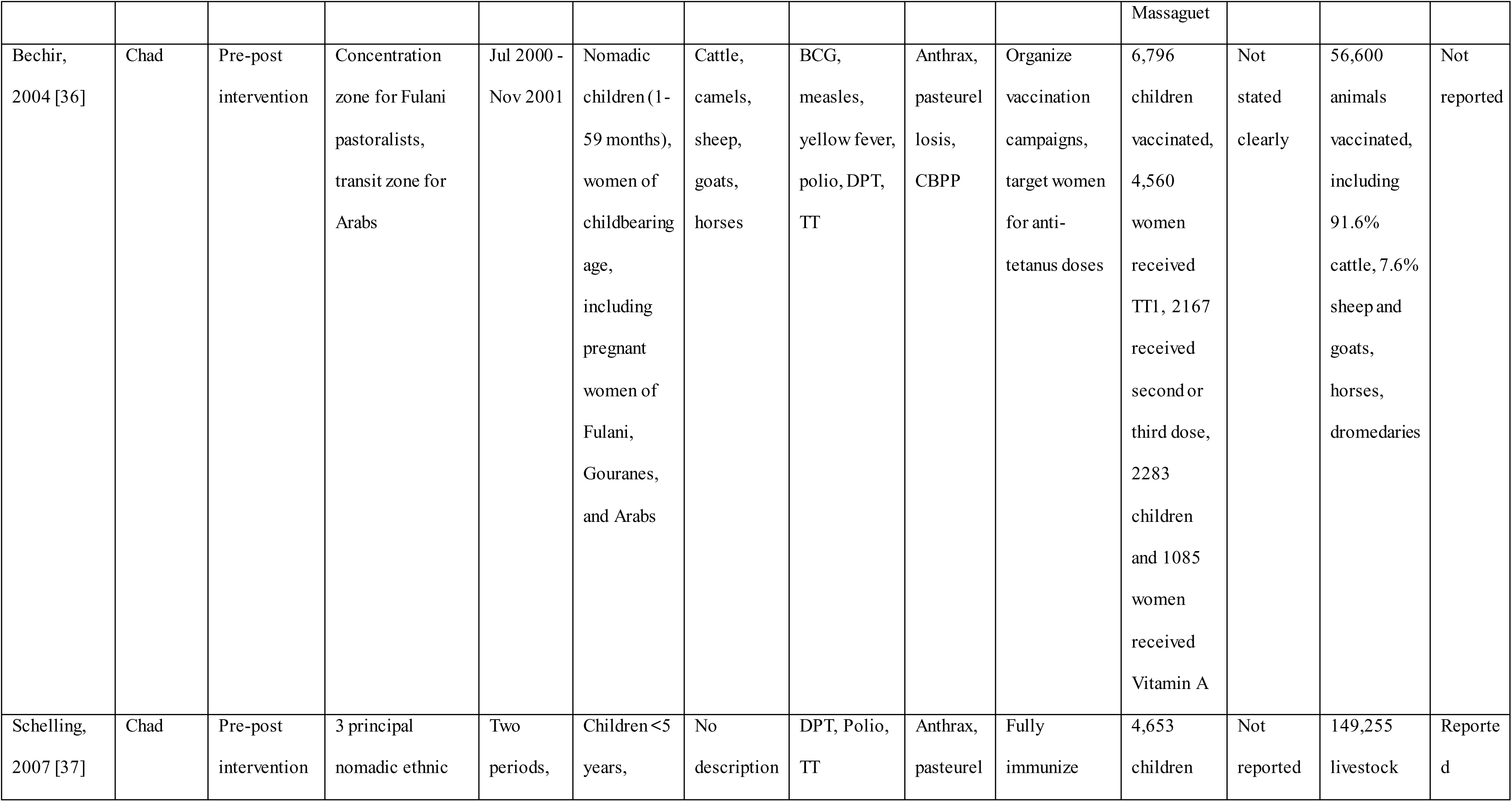
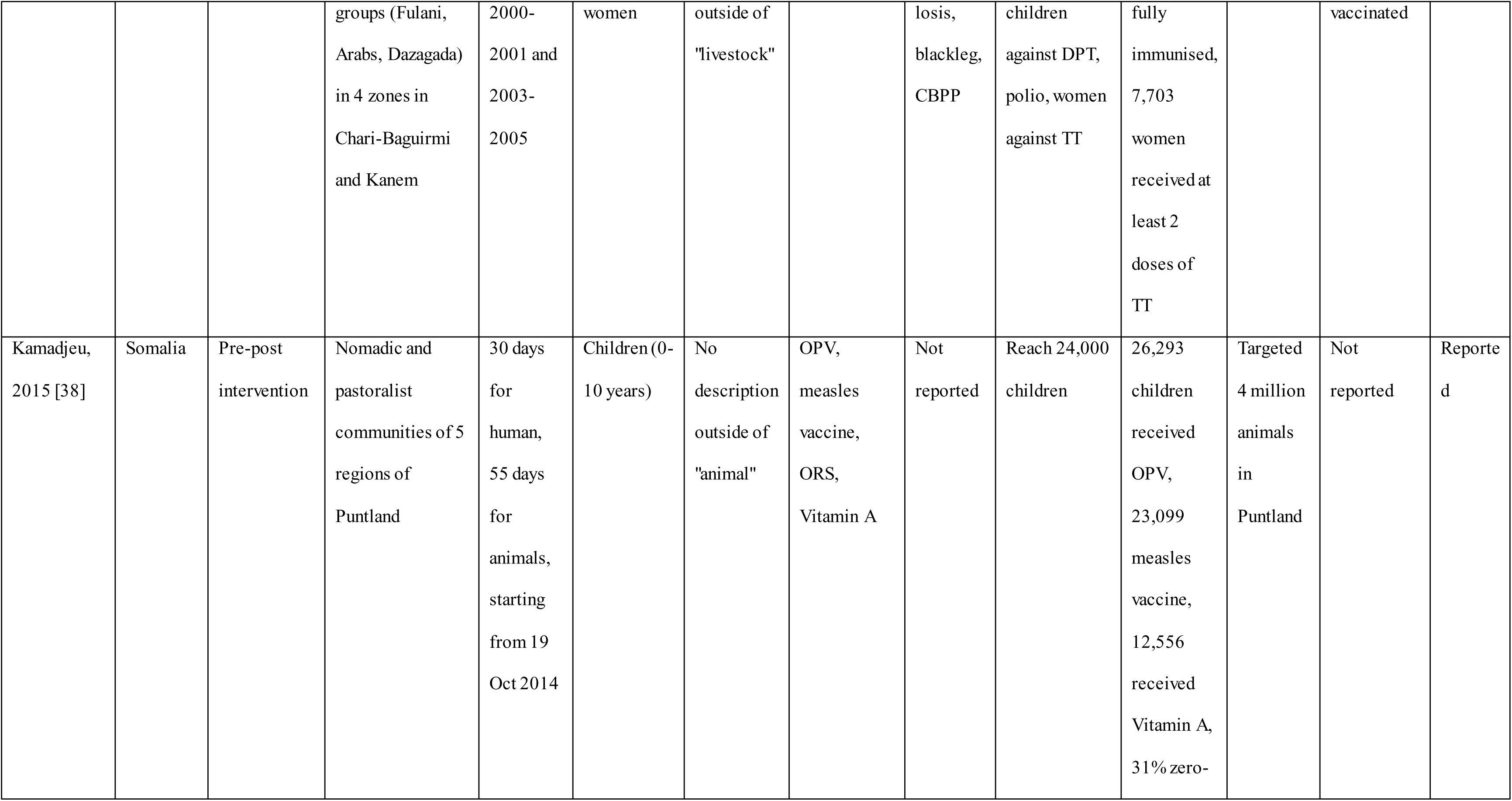
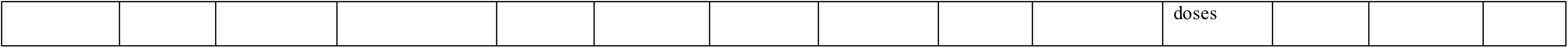
Studies reported in which joint human and animal vaccination was conducted to improve vaccination coverage among mobile pastoralist population heir animals in Africa.

### 3.3. Findings

The five records described activities aimed at conducting joint human and animal vaccination campaigns to improve vaccination coverage among the mobile pastoralist population and their animals. The study designs varied, with four using a pre-post intervention and one employing a community intervention trial with comparison districts [35].

#### Effect of the joint vaccination campaigns on human vaccination

Three records [34,36,37] focused on vaccinating children under five years and women of childbearing age, including pregnant women and two studies [35,38] focused on vaccinating children with the polio vaccine. The polio vaccine was used in the five records, with two studies conducted as campaigns to respond to wild poliovirus outbreaks in Chad [35] and Somalia [38]. The diphtheria, tetanus and pertussis (DTP)-containing vaccine was used in four studies [34,35,36,38]. The measles vaccine was used in three studies [35,36,37]. Tetanus toxoid vaccine (TT) was used in studies that included women of childbearing age [34,36,37]. Vitamin A was delivered as part of interventions in two records [35,38].

Four records reported changes in vaccination status in people based on the baseline results, which were presented in the form of vaccination coverage and/or the proportion of zero-dose children being vaccinated and the dropout rate [34,35,36,38]. Three records reported an increase in vaccination coverage; one reported increased coverage by 117%-176% among nomadic children, in addition to 5134 children vaccinated in an ad hoc campaign at the border between Chad and Cameroon [35]. A second study reported coverage increased from 22.7% to 80.1% among nomadic children in Nigeria [34], and the polio vaccination campaign exceeded its initial target by 9.6% [38].

A significant proportion of zero-dose and under-vaccinated children were identified in two and one records, respectively. ‘Zero-dose’ refers to children who have not received any vaccine dose, but Gavi, the Vaccine Alliance, links the term to not receiving the first DTP dose for operational reasons. [39]. In the Nigerian study, 24% and 41.6% of children never received the pentavalent and polio vaccines, respectively, and 30.5% of women who received TT received it for the first time [34]. Overall, 31% of all children under 10 years in the study from Somalia were vaccinated for the first time [38]. Under-vaccination was also reported during the joint vaccination campaigns in a study in Chad, which showed that approximately one-third of children (31%) had had three contacts with the vaccination team, and 48% of women had received a second or third dose of TT [36]. Two records reported that joint human and animal vaccination campaigns were conducted in response to polio outbreaks [35,38].

#### Effect of the joint vaccination campaigns on animal vaccination

Records reported substantial animal vaccination coverage during the joint campaigns. In Chad, 149,255 livestock were vaccinated, including 52,000 cattle fully immunised in Chari-Baguirmi and Kanem prefectures, with cattle constituting 91.6% of all vaccinated animals (approximately 56,600 total), while sheep and goats comprised 7.6%, with horses and camels making up the remainder [36,37]. In Northern Nigeria, vaccination coverage for 28,581 cattle increased from 41% to 61%, and 26 dogs were vaccinated against rabies [34]. Diseases targeted, when mentioned, included anthrax (*Bacillus anthracis*), blackleg (*Clostridium chauvoei*), pasteurellosis, and contagious bovine pleuropneumonia (CBPP; *Mycoplasma mycoides* subsp. *Mycoides*) for cattle, and rabies for dogs. The remaining two records [35,38] mentioned animal vaccination; however, they provided few or no details on species and diseases. Ndiaye et al. [35] noted the vaccination of ruminant livestock against anthrax and blackleg.

The value of animal vaccination was associated with several factors, including economics, zoonotic disease control, and serving as an entry point (platform) for human vaccination. It was stated that for nomadic pastoralists, livestock represents not only their primary livelihood but also their accumulated wealth, social status, and food security [37]. In addition, anthrax and rabies are zoonoses, and joint vaccination protects both animal and human health from these shared threats [37]. Another record stated that nomadic movements can spread animal diseases across wide geographic areas; targeted vaccination helps create buffer zones and reduce disease transmission during seasonal migrations [36]. Lastly, the high value placed on animal health makes veterinary services highly sought after, creating an effective platform for delivering human health interventions that might otherwise be refused or inaccessible [34,37].

#### Phases of implementing joint vaccination campaigns

Implementation of the vaccination campaigns was organised in three phases (Figure 2). The pre-vaccination or planning phase focused on coordinating with the local health ministries, ministries of livestock [34–38], international stakeholders such as UNICEF [35,36,37,38], WHO [35,38], FAO [38], US CDC [35], conducting a literature review [34], mapping nomadic movements [34,38], and engaging community leaders and volunteers [35,36,37,38]. During the implementation phase, teams, including vaccinators and community mobilisers, conducted multiple rounds of vaccinations [34–38], strategically timing campaigns to coincide with cattle crossings at known borders and seasonal settlement periods to maximise outreach [34–38]. In the post-vaccination phase, the effectiveness of the campaigns was assessed and reported. The costs of vaccination campaigns were reported in three studies [36–38]. Challenges such as high population mobility, logistical constraints, and difficulties with follow-up were identified [34–38].

**Figure 2:**
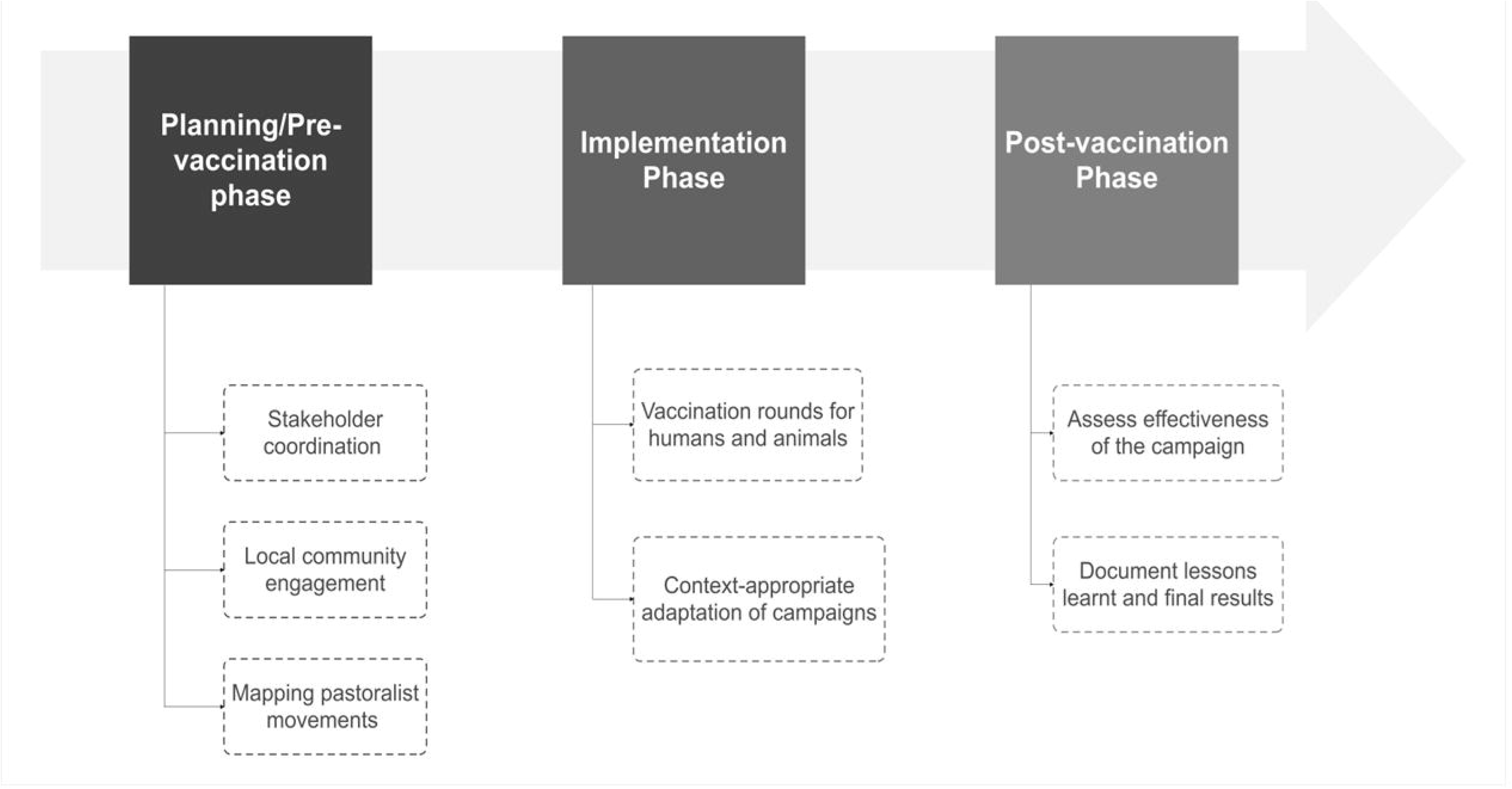
Phases of implementing joint campaigns for human and animal vaccinations

#### Enabling factors to implement joint vaccination campaigns

There were several key enabling factors reported among the studies. Political will was reported to be foundational to a system’s ability to prioritise the joint vaccination campaign. One record in Chad [35] highlighted the crucial role of national political support, noting that after a major outbreak in 2011, President Idriss Deby instructed regional governors to boost vaccination efforts, which in turn prompted the Ministry of Health’s Expanded Program on Immunization (EPI) to improve coverage among nomadic groups [35].

Building the joint vaccination campaign on existing regular veterinary services was one of the key factors reported in three records [35, 37, 38]. For example, one record in Chad noted that they collaborated with institutions including the Centre de Support en Santé International, a national centre specialising in rural health with many years of experience in projects directed at nomads and in integrating veterinary and human health services, in addition to a well-organised association of young and adult educated nomads [35]. Another record in Chad built upon existing compulsory animal vaccination [37]. A record in Somalia reported that the joint vaccination campaign was built upon the regular animal vaccination campaign, which is conducted by FAO twice a year to protect livestock vital to the livelihoods of Somali pastoralist and nomadic communities [38].

All studies reported a participatory approach throughout both the planning and implementation phases, with a strong emphasis on intersectoral collaboration and the active engagement of the community in all activities [35–38]. For example, a well-organised group of educated young and adult nomads in Chad was identified [35], and some members of this group were recruited and trained to conduct a census of children under five years old and pregnant women. Practical considerations, such as selecting appropriate seasons and corridors that aligned with the population’s needs, were also reported by all studies [35,36,37,38]. Three studies reported the importance of adapting interventions to ensure they were contextually acceptable [35,36,38]. Two of them [36,38] reported adapting the information, education, and communication (IEC) tools to align with the worldview of mobile pastoralists. For example, one study explained that in one IEC session, the importance of taking three doses of certain vaccines was illustrated using the analogy of building a tent in three stages: laying the foundation to represent the first dose, erecting the timber frame for the second, and laying the mats for the third. The tent can only offer protection once all three stages are completed [36].

One study [36] indicated that the intervention was adapted to ensure cultural acceptance, because individuals from different family groups refused to gather due to fears surrounding the "evil eye" affecting their livestock and children, which is widely believed among nomads in the Sahel region [5]. As a result, each group requested a vaccination centre near their respective camps or wells.

## 4. Discussion

Formal health services, including vaccination, overlook mobile pastoralists, including transhumant, semi-nomadic, and nomadic groups. Previous efforts to address this have not been systematically explored, especially regarding vaccination. This review of the literature on joint human and animal vaccination interventions for mobile pastoralists in Africa adds to our understanding of their implementation and effectiveness in reaching these underserved communities. The review identified enabling factors necessary for the successful implementation of joint human-animal vaccination interventions. These factors include the adoption of a culturally sensitive approach that improves accessibility and utilisation, logistical coordination, resource allocation, and community engagement in research activities.

Despite the small number of studies included in the analysis, the data obtained were extensive. However, it also highlights a significant gap in research focus on these vital populations and underscores the need for a deeper understanding of strategic entry points for human health interventions by recognising the social and economic value of livestock for these communities. Despite variations in study design, the collective findings suggest that integrating vaccination efforts can lead to increases in vaccine uptake among mobile pastoralists. However, these gains are tempered by the persistent challenge of high proportions of zero-dose children, highlighting the ongoing need to reach those who remain unvaccinated.

One of our key findings was the disproportionate attention given to different geographic regions within the broader context of mobile pastoralists in Africa and the methodological approaches employed. Specifically, four studies were conducted in the Sahel region, with Chad contributing three studies that documented joint human and animal vaccination efforts. Mobile pastoralists have long been present across multiple countries in the Sahel and Horn of Africa regions [40]. However, due to limited documentation, it is possible that some joint vaccination campaigns may have occurred without formal records. For instance, a qualitative study in Sudan reported that non-regular co-delivery of vaccinations for both humans and animals has been conducted, facilitated through intersectoral collaboration between the Ministry of Health and the Ministry of Animal Resources [5]. We suggest increasing documentation of joint human and animal campaign activities to gather insights into the best approaches and practices among these often-overlooked yet vulnerable groups, and to demonstrate how these campaigns operate in different context-specific settings.

Most of the studies identified in this review employed pre- and post-approaches, suggesting that joint human and animal vaccination efforts were primarily conducted as supplementary immunisation activities (SIAs) or campaigns rather than as methodologically designed studies. These methods were likely to be helpful in the context of nomadic populations, particularly in overcoming challenges such as insufficient funding, difficulties in monitoring participant compliance, and participants relocating to different areas. This also highlights the scarcity of research among pastoralists and the limited diversity in study design, as evidenced by the presence of only one study featuring interventional comparison groups.

Although this review described studies that delivered joint vaccination, a promising One Health approach that supports the non-hierarchical worldviews of nomadic pastoral people, it also highlighted persistent shortcomings in joint human-animal health strategies [27, 28]. Despite purporting to prioritise equitable animal and human health, these approaches often still place human health at the forefront, thereby overlooking the non-hierarchical nature of pastoralists’ values. Literature highlights critical gaps between the ideals of One Health and its practical application, in which human health outcomes often overshadow those of animals [27]. This imbalance is also evident in our findings, which emphasise human vaccination outcomes while giving minimal or no consideration to animal health outcomes. This overlooks the vital role of animals in pastoralist communities as sources of livelihood and social identity, despite animal health services serving as an entry point for these communities and an established platform to support joint vaccination [18,25,26]. The diminished focus on animal health might be due to vaccination campaigns primarily targeting nomadic humans with a focus on polio elimination in Chad, Nigeria, and Somalia, where wild poliovirus cases were prevalent. The polio vaccine was administered in all the studies identified in this review. Notably, the last reported cases of polio in Chad and Somalia were reported among nomadic communities during the period of joint vaccination efforts [7–9]. For a genuinely integrated One Health approach to take shape, it is essential to equitably recognise and operationalise the interdependence between human and animal health, especially in settings where animals are integral to community well-being.

Our findings suggest that vaccination status, particularly among nomadic children and women of reproductive age, improved after implementing joint human and animal vaccination strategies. Despite high dropout rates, these efforts reduced the proportion of zero-dose children who had never received any vaccinations, indicating that joint campaigns could be an effective approach for vaccination delivery in nomadic and mobile communities. The most recent documentation of these joint strategies was recorded in 2016 [34], before the launch of the Immunization Agenda 2030 (IA2030), introduced by various stakeholders in immunisation, including WHO, UNICEF, and Gavi, the Vaccine Alliance. This agenda, which spans the period from 2021 to 2030, aims to enhance immunisation "coverage and equity," ensuring that everyone receives protection through vaccination and reducing the number of zero-dose children by 50% by 2030 [41]. If implemented effectively in the future, these joint strategies could help achieve the objectives outlined in IA 2030.

Our study only identified five studies for inclusion, indicating a scarcity of published studies and grey literature addressing the benefits and challenges of joint vaccination delivery. This is likely an incomplete picture of the subject. Further insights might be available from unpublished reports or ad-hoc local initiatives that have never been described. Pastoralist communities differ in cultural practices, mobility patterns, geographical settings, and interactions with state or traditional systems. This variability means that a strategy that is effective in one context may not be effective in another. Developing universally replicable recommendations is challenging, and context-specific strategies continue to be necessary. To achieve this, further investigation into joint delivery should be conducted, using the findings of this review as a foundation. The goal of IA2030 is to leave no one behind [41]. With the substantial reductions in Official Development Assistance (ODA) by major donor governments in 2025, innovative approaches are needed to reach these mobile pastoralists with vaccination services. The One Health approach is currently a missed opportunity and has the potential to realise the aspiration of IA2030 by considering the context-specific realities of mobile pastoralists, including non-hierarchical thinking and a holistic, non-anthropocentric perspective. Enhanced collaboration is necessary to reduce the cost at the country level among the respective ministries of health and livestock in addition to the Quadripartite organisations at the regional and country levels- the WHO, the Food and Agriculture Organization of the United Nations (FAO), the United Nations Environment Programme (UNEP) and the World Organisation for Animal Health (WOAH).

## Conclusion

Our review revealed that joint vaccination of animals and people in nomadic pastoralist populations within a limited region in Africa resulted in improved vaccination rates for several critical childhood and animal diseases, including zoonoses. It provided insights into employing a pro-equitable approach to engaging mobile pastoralist communities that face various forms of marginalisation. This is crucial when implementing responsive interventions that respect their mobility and lifestyle identity, ensuring services are provided through an equitable rather than a restrictive lens. However, considering the last reported joint interventions and the evolving geopolitical dynamics in both the Horn of Africa and the Sahel, including the rising number and complexity of conflicts, as well as their intersection with climate change, more research is essential to assess the feasibility and cost-effectiveness of these approaches. Whilst joint vaccination campaigns appear to improve human vaccine uptake, more information is needed to evaluate the potential benefits that could be gained by attention to animal vaccination and improved animal health outcomes that could further improve non-vaccine-related human health outcomes

Due to the frequent transboundary movements of mobile pastoralists within the Horn of Africa and the Sahel region, it is imperative to establish a comprehensive regional joint vaccination policy and strategy under the auspices of the African Union. These joint policies and strategies will help put these into practice for these significant populations. They should involve regional organisations, including the Intergovernmental Authority on Development (IGAD), which can support mobile pastoralists in the Horn of Africa, and the Economic Community of West African States (ECOWAS), which can coordinate these efforts in the Sahel.

## Data Availability

All data are available in the manuscript and supporting information files.

