## Supplementary 1 for "Joint human and animal vaccination strategies to reach mobile pastoralists and their livestock in Africa: A scoping review"

**Appendix A. Ovid Medline search strategy**

1	exp Immunization/

2	exp Immunization Programs/

3	exp Vaccines/

4	(immuniz$ or immunis$ or inoculat$ or vaccin$).tw,kw.

5	1 or 2 or 3 or 4

6	exp "Transients and Migrants"/

7	(nomad$ or semi-nomad$ or seminomad$).tw,kw.

8	pastoralis$.tw,kw.

9	((itinerant$ or transient$ or mobile or moving) adj4 (migrant$ or farmer$ or herder$ or herdsm$ or herdswom$ or agrarian$ or agricultural$ or grazier$ or population$)).tw,kw.

10	transhuman$.tw,kw.

11	fulani$.tw,kw.

12	afar$.tw,kw.

13	maasai$.tw,kw.

14	borana$.tw,kw.

15	bedouin$.tw,kw.

16	turkana$.tw,kw.

17	tuareg$.tw,kw.

18	6 or 7 or 8 or 9 or 10 or 11 or 12 or 13 or 14 or 15 or 16 or 17

19	5 and 18

20	exp Africa/

21	africa$.tw,kw.

22	exp Algeria/

23	algeria$.tw,kw.

24	exp Angola/

25	angola$.tw,kw.

26	exp Benin/

27	benin$.tw,kw.

28	exp Botswana/

29	botswana$.tw,kw.

30	exp Burkina Faso/

31	(burkina adj1 faso$).tw,kw.

32	burkinabe$.tw,kw.

33	exp Burundi/

34	burundi$.tw,kw.

35	exp Cabo Verde/

36	((cape or cabo) adj1 verde$).tw,kw.

37	exp Cameroon/

38	Cameroon$.tw,kw.

39	exp Central African Republic/

40	(Central adj1 African$).tw,kw.

41	exp Chad/

42	Chad$.tw,kw.

43	exp Comoros/

44	Comor$.tw,kw.

45	exp "Democratic Republic of the Congo"/

46	(Democratic adj1 Republic$ adj3 Congo$).tw,kw.

47	congoles$.tw,kw.

48	exp Congo/

49	(Republic adj3 Congo$).tw,kw.

50	exp Cote d'Ivoire/

51	(Cote adj1 d'Ivoire$).tw,kw.

52	ivorian$.tw,kw.

53	exp Djibouti/

54	Djibouti$.tw,kw.

55	exp Egypt/

56	Egypt$.tw,kw.

57	exp Equatorial Guinea/

58	(Equatorial adj1 Guinea$).tw,kw.

59	exp Eritrea/

60	Eritrea$.tw,kw.

61	exp Eswatini/

62	Eswatini$.tw,kw.

63	swazi$.tw,kw.

64	exp Ethiopia/

65	Ethiopia$.tw,kw.

66	exp Gabon/

67	Gabon$.tw,kw.

68	exp Gambia/

69	Gambia$.tw,kw.

70	exp Ghana/

71	Ghan$.tw,kw.

72	exp Guinea/

73	Guinea$.tw,kw.

74	exp Guinea-Bissau/

75	Guinea-Bissau$.tw,kw.

76	exp Kenya/

77	Kenya$.tw,kw.

78	exp Lesotho/

79	Lesotho$.tw,kw.

80	basotho$.tw,kw.

81	exp Liberia/

82	Liberia$.tw,kw.

83	exp Libya/

84	Libya$.tw,kw.

85	exp Madagascar/

86	Madagasca$.tw,kw.

87	exp Malawi/

88	Malawi$.tw,kw.

89	exp Mali/

90	Mali$.tw,kw.

91	exp Mauritania/

92	Mauritania$.tw,kw.

93	exp Mauritius/

94	Mauriti$.tw,kw.

95	exp Morocco/

96	Morocco$.tw,kw.

97	exp Mozambique/

98	Mozambi$.tw,kw.

99	exp Namibia/

100	Namibia$.tw,kw.

101	exp Niger/

102	Niger$.tw,kw.

103	exp Nigeria/

104	exp Rwanda/

105	Rwanda$.tw,kw.

106	exp "Sao Tome and Principe"/

107	(Sao adj1 Tome adj2 Principe$).tw,kw.

108	(sao adj1 tomean$).tw,kw.

109	exp Senegal/

110	Senegal$.tw,kw.

111	exp Seychelles/

112	Seychell$.tw,kw.

113	exp Sierra Leone/

114	(Sierra adj1 Leone$).tw,kw.

115	exp Somalia/

116	Somali$.tw,kw.

117	exp South Africa/

118	(South adj1 Africa$).tw,kw.

119	exp South Sudan/

120	(South adj1 Sudan$).tw,kw.

121	exp Sudan/

122	Sudan$.tw,kw.

123	exp Tanzania/

124	Tanzania$.tw,kw.

125	exp Togo/

126	Togo$.tw,kw.

127	exp Tunisia/

128	Tunisia$.tw,kw.

129	exp Uganda/

130	Uganda$.tw,kw.

131	exp Zambia/

132	Zambia$.tw,kw.

133	exp Zimbabwe/

134	Zimbabwe$.tw,kw.

135	sahara$.tw,kw.

136	sahel$.tw,kw.

137	20 or 21 or 22 or 23 or 24 or 25 or 26 or 27 or 28 or 29 or 30 or 31 or 32 or 33 or 34 or 35 or 36 or 37 or 38 or 39 or 40 or 41 or 42 or 43 or 44 or 45 or 46 or 47 or 48 or 49 or 50 or 51 or 52 or 53 or 54 or 55 or 56 or 57 or 58 or 59 or 60 or 61 or 62 or 63 or 64 or 65 or 66 or 67 or 68 or 69 or 70 or 71 or 72 or 73 or 74 or 75 or 76 or 77 or 78 or 79 or 80 or 81 or 82 or 83 or 84 or 85 or 86 or 87 or 88 or 89 or 90 or 91 or 92 or 93 or 94 or 95 or 96 or 97 or 98 or 99 or 100 or 101 or 102 or 103 or 104 or 105 or 106 or 107 or 108 or 109 or 110 or 111 or 112 or 113 or 114 or 115 or 116 or 117 or 118 or 119 or 120 or 121 or 122 or 123 or 124 or 125 or 126 or 127 or 128 or 129 or 130 or 131 or 132 or 133 or 134 or 135 or 136

138	19 and 137
